# Neuromotor Prosthetic to Treat Stroke-Related Paresis

**DOI:** 10.1101/2021.02.03.21250720

**Authors:** Mijail D. Serruya, Alessandro Napoli, Nicholas Satterthwaite, Joseph Kardine, Joseph McCoy, Namrata Grampurohit, Kiran Talekar, Devon Middleton, Feroze Mohamed, Michael Kogan, Ashwini Sharan, Chengyuan Wu, Robert Rosenwasser

## Abstract

**Background:** Functional recovery of independent arm movement typically plateaus within six months following a stroke, leaving chronic motor deficits. This feasibility study tested whether a wearable, powered exoskeletal orthosis, driven by a percutaneous, implanted brain–computer interface (BCI), using the activity of neurons in the precentral gyrus in the affected cortical hemisphere, could restore voluntary upper extremity function in a person with chronic hemiparetic subsequent to a cerebral hemispheric stroke of subcortical gray and white matter and cortical gray matter.

**Methods:** One person with chronic hemiparetic stroke with upper-limb motor impairment used a powered elbow-wrist-hand orthosis that opened and closed the affected hand using cortical activity, recorded from four 64-channel microelectrode arrays implanted in the ipsilesional precentral gyrus, based on decoding of spiking patterns and high frequency field potentials generated by imagined hand movements using technology and decoding methods used for people with other causes of paralysis. The system was evaluated in a home setting daily for 12 weeks.

**Results:** Robust single unit activity, modulating with attempted or imagined movement, was present throughout the precentral gyrus areas. The participant was able to acquire voluntary control over a hand-orthosis BCI, with a score of 10 points on the Action Research Arm Test (out of 53) using the BCI, compared to 0 without any device, and 5 using myoelectric control. Orthosis-powered hand-opening was faster with BCI control compared to myoelectric control, on a standardized object-movement task.

**Conclusions:** The findings demonstrate the therapeutic potential of an implantable BCI system coupled to a brace to “electrically bypass” the stroke and promote neurally driven limb function. The participant’s ability to rapidly acquire voluntary control over otherwise paralyzed hand opening, more than 18 months after a subcortical stroke, lays the foundation for a fully implanted movement restoration system.

## Inroduction

Stroke is a leading cause of disability ^1^ with a global prevalence of over 42 million people in 2015 ^2^, affecting over four million adults in the United States alone with 800,000 new cases per year ^3^. Stroke leads to permanent motor disabilities in 80% of cases ^4^, and half of stroke survivors require long term care. Brain computer interface (BCI) technologies offer a potential solution to restore functional independence and improve health in people living with its effects. In the past decade, intracortical BCI technology has continued to advance, with multiple groups demonstrating the safety and efficacy of this approach to derive control signals ^5–7^ to restore communication and control. In parallel, wearable robotic orthosis technology can benefit patients with weakened limbs^8,9^. This single-patient pilot clinical trial sought to prove that a commercially available powered arm orthosis could be linked to the cerebral cortex in an adult with the most common form of chronic stroke. A direct path from the brain’s motor centers to the orthotic could reanimate a paralyzed limb to enable useful hand and arm function.

Several signal sources have been coopted to provide commands to move paralyzed limbs. Electromyographic (EMG) control of a powered orthosis or functional electrical stimulation (FES) of muscles, has proven problematic either because users could not generate sufficient or reliable activity to provide a good control signal, or because voluntary activation of those recorded muscles (that were intended to generate the command) was opposed by the stimulator’s effects ^10^. Contralaterally controlled electrical stimulation-where activity from the unaffected arm triggers stimulation on the paretic arm- is a useful therapeutic intervention to improve function in the weaker limb^11^ but it is not clear how this unnatural command source could be generalized to continuously-worn devices that enable independent arm movements. Several groups have explored scalp EEG, which is closer to the command’s origins, to derive control signals to drive robotic braces, and in one case, FES ^8,12–14^. While using EEG-derived signals may be promising for rehabilitation therapy, it would not be feasible for daily independent function because skin sweat and hair can cause impedances to fluctuate, compromising signal quality. Daily application of even a subset of contacts to the same skin sites can lead to skin breakdown and cellulitis. Further, EEG signals are limited in the commands that can be easily and reliably derived from the available signal. By contrast, intracortical interfaces offer a rich sources of high resolution, multidimensional control signals, since it is the origin of such signals in healthy adults, in non-human primates and in people with spinal or brainstem disorders ^7,15,16^.

While the vast majority of strokes involve cerebral white matter and even direct parenchymal damage, intracortical neuromotor prosthetics have not been tested in people with strokes above the mesencephalon. It is not known whether motor cortex remains a reliable signal source in this large population. A proof-of-concept that a brain-computer interface, based on micro-electrode arrays implanted in intact cortex above a subcortical stroke, could restore behaviorally useful independent, voluntary movement, could lead to the development of a fully implantable medical device that, in principle, could reverse the motor deficits caused by stroke.

## Methods

Approval for this study was granted by the US Food and Drug Administration (Investigational Device Exemption) and the Thomas Jefferson University Institutional Review Board. The participant described in this report has provided permission for photographs, videos and portions of his protected health information to be published for scientific and educational purposes. After completion of informed consent, medical and surgical screening procedures, two MultiPorts (Blackrock Microsystems, UT), each comprising two 8×8 platinum tipped microelectrode arrays tethered to a titanium pedestal connector, were implanted into the cortex of the precentral gyrus using a pneumatic insertion technique^17,18^. Details of the human surgical procedure are in preparation for publication and followed other similar studies. Trial selection criteria are available online (see Clinicaltrials.gov, NCT03913286). The trial was designed with the implantation phase to last a maximum of three months (Fig. 1).

**Figure 1.**
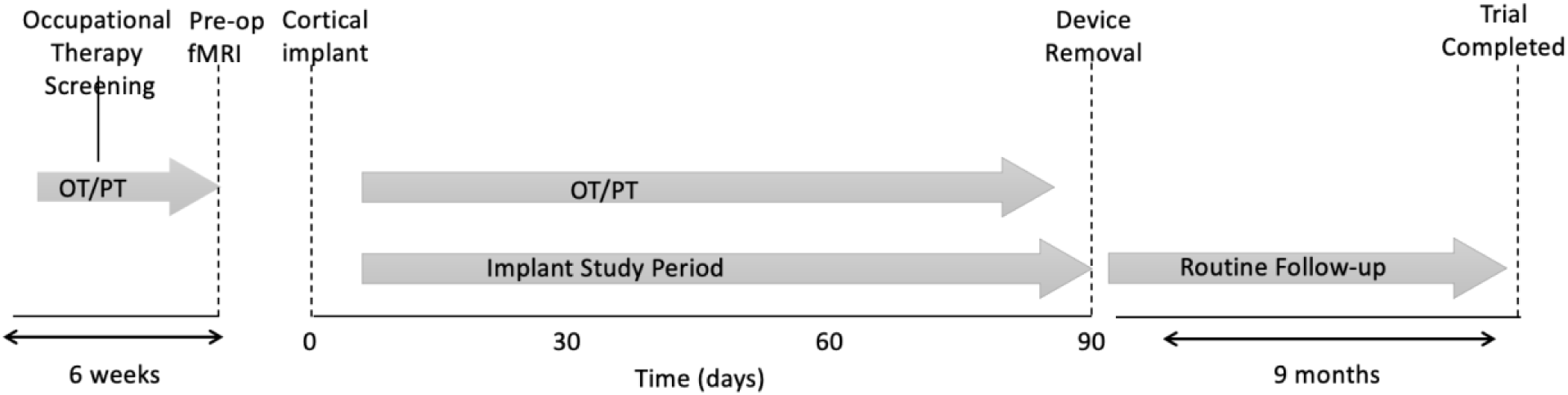
The clinical trial timeline.

### Participant

The participant was a right-handed male who experienced right hemispheric stroke, manifest as acute onset dense left hemiparesis and expressive aphasia, at which time he was age between ages 35 to 40. Due to unknown time of onset and hypertension at presentation, the participant was not a candidate for thrombolysis. CT angiogram showed occlusion of the right posterior cerebral artery and high-grade stenosis of the left posterior cerebral artery in the proximal P2 segment. MRI of the brain showed acute infarcts in the right basal ganglia/corona radiata and right occipital lobe. He was started on dual antiplatelet therapy for 3 weeks and then was transitioned to aspirin 81 mg once daily, along with atorvastatin and anti-hypertensives. He had left-sided hemiparesis, dysphagia, left homonymous hemianopsia and dense left visual neglect and was transferred for inpatient rehabilitation. Over a period of three months, aphasia and dysphagia resolved and he learned to ambulate independently, albeit with a persistent left foot drop. Neuroimaging showed evidence of multi-focal strokes, and prior silent strokes. The participant had previously been in good health and did not have any known stroke risk factors such as diabetes or smoking. There was a history of loud snoring and the participant had not been evaluated for obstructive sleep apnea. Transthoracic and transesophageal echocardiography were normal as were serial hypercoagulability panels; the participant was adopted and the biological family history unknown. The participant was deemed to have had embolic strokes of unknown source. Although serial electrocardiography since the stroke was normal, the participant is being scheduled for a loop recorder to survey for possible paroxysmal atrial fibrillation. The participant had learning disabilities and was presumed to have had mild cognitive impairment prior to the stroke. Screening formal neuropsychological testing identified neurocognitive problems (full scale IQ 59) and also concluded that the participant remained fully capable to provide proper informed consent and to participate in this trial, meeting its demands and requirements. The participant provided both verbal and written informed consent, both to participate in the trial and to share his identifying information with the public. He had been working full time at the time of the stroke and has been unable to return to work since the stroke.

### Pre-operative fMRI

The participant underwent MRI on a 3T Philips Ingenia MRI scanner. A 1mm isotropic 3-D T2 FLAIR was obtained for structural localization. A single-shot echoplanar gradient echo imaging sequence with 80 volumes, repetition time (TR) = 2 s, echo time (TE) = 25 msec, voxel size = 3 × 3 mm2, slice thickness = 3 mm, axial slices = 37. The participant was asked to visualize movements of his paretic left hand during the MRI. Each motor trial consisted of a block design featuring a 20 s rest block and a 20 s active block repeated. This block design was repeated between 4 times for a total of 240 s scans. Visual stimuli comprised a 20 s video depicting a 3D modeled limb at rest, followed by a 20 s video of the limb performing the desired task. Motor tasks included repeated hand open/clench or arm extension elbow, and were either “active” (participant performed or attempted to perform motion) or “passive” (physician manually moved participant’s arm). In active tasks, the participant was instructed to follow the movements in the video or concentrate on following for the paretic limb. Task prioritization was based on pre-exam training of the participant’s capabilities and examination of BOLD activation observed during the scan. Post processing including motion correction, smoothing, and general linear model estimation performed using SPM software (www.fil.ion.ucl.ac.uk/spm) and Nordic brain EX software (NordicNeuroLab, Bergen, Norway). Statistical maps were overlaid on the 3D T2 FLAIR image for visualization of activation.

### Cortimo system

‘Cortimo’ is the designation provided to the FDA to represent the overall system (Fig. 2) that comprised two percutaneous Multiports (Blackrock Microsystems), each in turn having two multi-electrode array sensors, the cabling, amplifiers, software and the powered MyoPro orthosis. Each sensor is an 8×8 array of silicon microelectrodes that protrude 1.5 mm from a 3.3 × 3.3-mm platform. At manufacture, electrodes had an impedance ranging between 70 KOhm and 340 KOhm. The arrays were implanted onto the surface of the MI arm/hand region guided by the pre-operative fMRI; with electrodes penetrating into the cortex to attempt to record neurons in layer V. Recorded electrical signals pass externally through a Ti percutaneous connector secured to the skull. Cabling attached to the connector during recording sessions routes signals to external amplifiers and a computer that process the signals and convert them into different outputs, such as servo motor position of the MyoPro brace or screen position of a neural cursor. Currently, this system must be set up and managed by an experienced technician.

**Figure 2.**
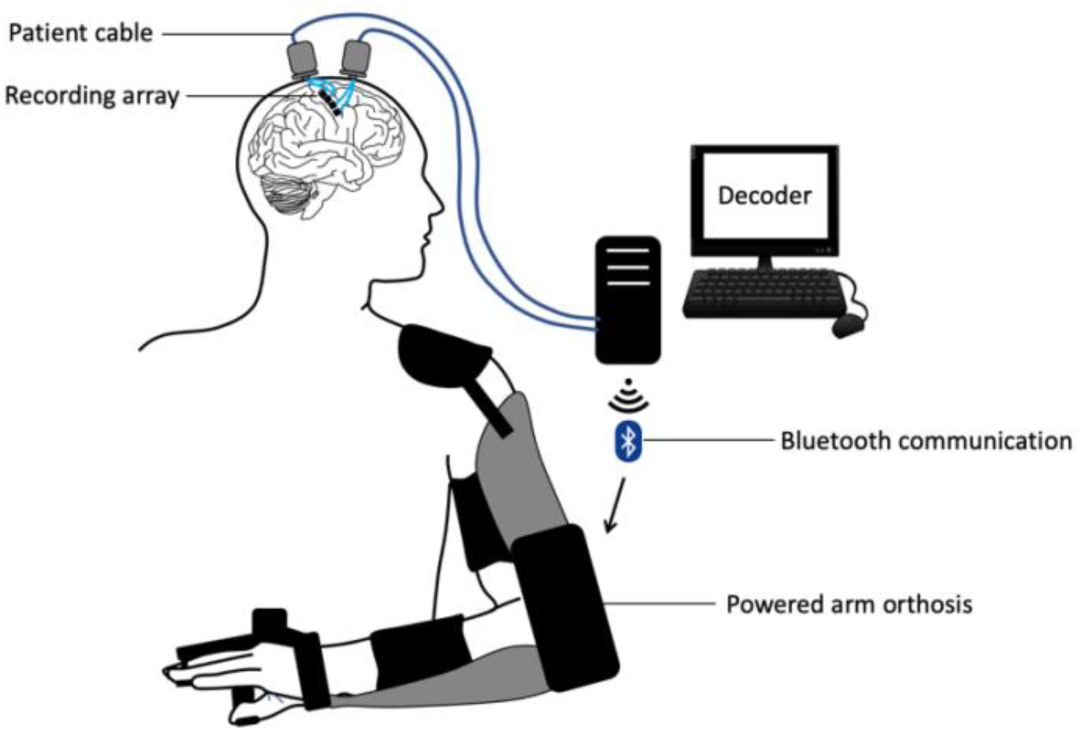
The Cortimo system. The overall device comprises two Multiports, each with two 8×8 microelectrode arrays, patient cables linked to external amplifiers, a decoding computer and a wearable, powered arm orthosis.

### MyoPro brace

The MyoPro (Myomo, Inc, Cambridge, MA) is an FDA-cleared myoelectric powered arm orthosis designed to support a paretic arm^9^.The rigid brace incorporates metal contacts attached to soft straps that can be adjusted such that contacts rest on the biceps and triceps proximally, and on wrist flexors and extensors distally, on the paretic upper extremity. The sensors continuously record the root mean square of underlying muscle activity. Thresholds are manually set such that signals exceeding them will trigger one of the MyoPro motors. Because the participant retained residual elbow flexion and extension strength, the motor at the elbow was set up such that biceps activation triggered elbow flexion and triceps activation triggered elbow extension. For hand opening, the MyoPro was set up to either use myoelectric control, or to use BCI-based control. Since the participant was unable to voluntarily extend the wrist or open the fingers, the myoelectric mode was set up such that the default state was with the hand open, and it would only be closed by activating enough wrist flexor activity.

### Recording sessions

Research sessions were scheduled five days per week at a temporary residence, adjacent to the hospital, provided to the participant. Sessions could be cancelled or ended early at the participant’s request. Sessions would commence with neural recording and spike discrimination. While initial sessions included filter building and structured clinical end-point (cursor control) trials, in the final month of the trial, “training-less” algorithms were used with the participant proceeding directly to BCI-controlled hand action once patient cables were connected. Performance of computer tasks, orthosis control and occupational therapy exercises followed. The electrodes and neural signals selected immediately before filter building remained constant for any given session’s orthosis control trials.

### Decoder Filter Building

Units were extracted using an automatic thresholding approach based for each electrode channel, based on Root Mean Squared multipliers^16^. For each session, single and multiunit data or high frequency (100-1000 Hz) local field potentials derived from multiple channels (20-30) were used to create a linear filter to convert these real-time multidimensional neural features into either a one or a two-dimensional (position or velocity) output signal. Motor activity and motor imagery approaches were tested for filter building, including imagining opening and closing the paretic hand, passively flexing and extending the elbow, passively opening and closing the hand, and observing a computer cursor displayed on a monitor moving up and down without any specific instruction. Training data for building the linear filter were collected with the participant gazing at a screen where a target cursor was moved slowly up and down for one minute (5 seconds to go from the top to the bottom of the screen or vice versa, at 20º visual angle). After this preliminary filter was built, a new 1-minute re-training session was performed, this time the manually controlled target cursor was accompanied by a prediction cursor that was neurally controlled by the participant. Using this additional training set, a second filter was built and then tested on a simple target acquisition game in which the y-position of the predicted output was discretized into zones such that positions on the upper part of the screen would cause an animation sprite to move up by a fixed distance (1 cm), and positions on the lower part of the screen would cause the sprite to move down by the same fixed distance.

### BCI Orthosis use

The discrete output was then used to control the aperture of the hand via the MyoPro’s hand brace motor. The up-down mapping on the screen was translated into closed-open positions of the hand. The participant then performed a series of functional tasks including grasping and then dropping an object, the Action Research Arm Test^19^, and a variation on Jebsen Taylor item moving test^20^. These were tested with both the participant seated and standing.

### “Training-less” Mapping

When the participant would attempt to overpower the orthosis motors with residual finger flexion strength, a novel ‘training-less’ approach was deployed in which a rolling 1-second baseline of the LFPs signals was used to calculate spectral power in the high gamma band (100-500 Hz). Namely, 1-second long LFP continuous voltages were used for computing the average spectral density estimation in the frequency band 100-500 Hz, using non-overlapping frequency bins with a 50Hz width. Spectral density was computed using the Matlab periodogram method. Values were updated every 500 ms, using 1-second-long rolling windows with 50% overlap. Real-time spectral features derived from the 20 most neuromodulated channels were averaged across channels to produce a single high gamma band value for each 500 ms software update. Orthosis hand-closure would be triggered by an increase in this mean spectral power from the resting baseline ranging between 0.5 and 3 V^2^/Hz to values greater than 10 V^2^/Hz, where real-time values above this threshold would make the hand motor close.

### Concomitant Occupational and Physical Therapy

Since being discharged from acute rehabilitation 60 days after the initial stroke, the participant enrolled in outpatient physical and occupational therapy. Prior to the device implantation, the participant completed a six-week course of occupational therapy screening phase. Following device implantation, the participant continued occupational therapy, twice per week, and physical therapy, once per week. Occupational therapy focused on postural training while seated and walking, donning and doffing the MyoPro, and using the MyoPro for functional activities. Timed functional electrical stimulation (e.g., pincer grasp programs; XCite, Restorative Therapies) and vibration therapy were used for spasticity management. Physical therapy exercises included scapular mobilization, progressive range of motion, weight bearing, forced use with game-related activities to encourage left UE volitional control, and aerobic endurance exercise.

## Results

The participant underwent intracortical implantation in autumn of 2020 and explantation three months later on January 2021, in accordance with the intended 3-month duration of the trial. Over the course of the study, the participant had three minor, and one serious, device-related adverse events, all of which were treated, resolved, and reported to the governing regulatory bodies. The serious adverse event was the development of a scalp infection at the left pedestal site one week prior to the device removal date, despite a regimen of topical antibiotics and regular cleaning. This infection was anticipated and was described as a potential risk in the informed consent form and consent interview. The left pedestal site had posed a challenge since the time of the initial surgery as it was not possible to exactly re-approximate the skin flap leaving the base of the pedestal exposed. This area was protected and granulated and grew new skin. The participant was afebrile and asymptomatic, and the infection was detected only by close visual inspection. The participant was treated with twice daily antibiotic for the 7 days prior to the device removal. Pedestal site skin cultures taken at device removal revealed pansensitive *staphylococcus lugdunesis* and *staphylococcus capitis*, and yeast, and appropriate antimicrobial treatment was provided. No organisms grew from cultures taken of adjacent bone. The only macroscopic evidence of infection at device removal was a small area (∼ 2 cm^3^) of erythema and friable tissue at the skin adjacent to the right pedestal. The participant was discharged home. The participant remains in the Cortimo trial for ongoing neurosurgical follow-up and surveillance, and to track any further performance improvements in myoelectric MyoPro use with ongoing outpatient occupational therapy.

### Preoperative anatomic and functional neuroimaging

Preoperative imaging revealed the old infarct in right lentiform nucleus and adjacent white matter including corona radiata and a portion of the posterior limb of the internal capsule, along with a large old right PCA infarct, progressed since the acute stroke imaging MRI from 2019 (Fig. 3). In addition, a small region of bandlike signal abnormality involving subcortical white matter and medial aspect of hand knob region of right precentral gyrus was identified, likely reflecting retrograde neuronal degeneration. On DTI, there was extensive loss of fractional anisotropy in the region of right corticospinal tract from old infarct. The “imagined” left hand motor paradigm and passive motor paradigm were diagnostic with good concordance. Subsequent to hypercapnia challenge, a BOLD signal was evident at the precentral gyrus. On the “imagined” left hand motor paradigm, activation was noted in the expected location along central sulcus involving lateral aspect of the hand knob region of the precentral gyrus and the adjacent portion of postcentral gyrus (Fig. 3c). On the passive left elbow motor paradigm, activation was seen along central sulcus which shows good concordance with the “imagined” motor task as discussed with a slightly more posterior and superior extension of activation reflecting the prominent sensory component of this passive motor paradigm. A 3D brain model was printed using the 3D FLAIR sequence to allow for 3D visualization of the surgical field for more accurate pre-operative planning (Fig. 3d).

**Figure 3.**
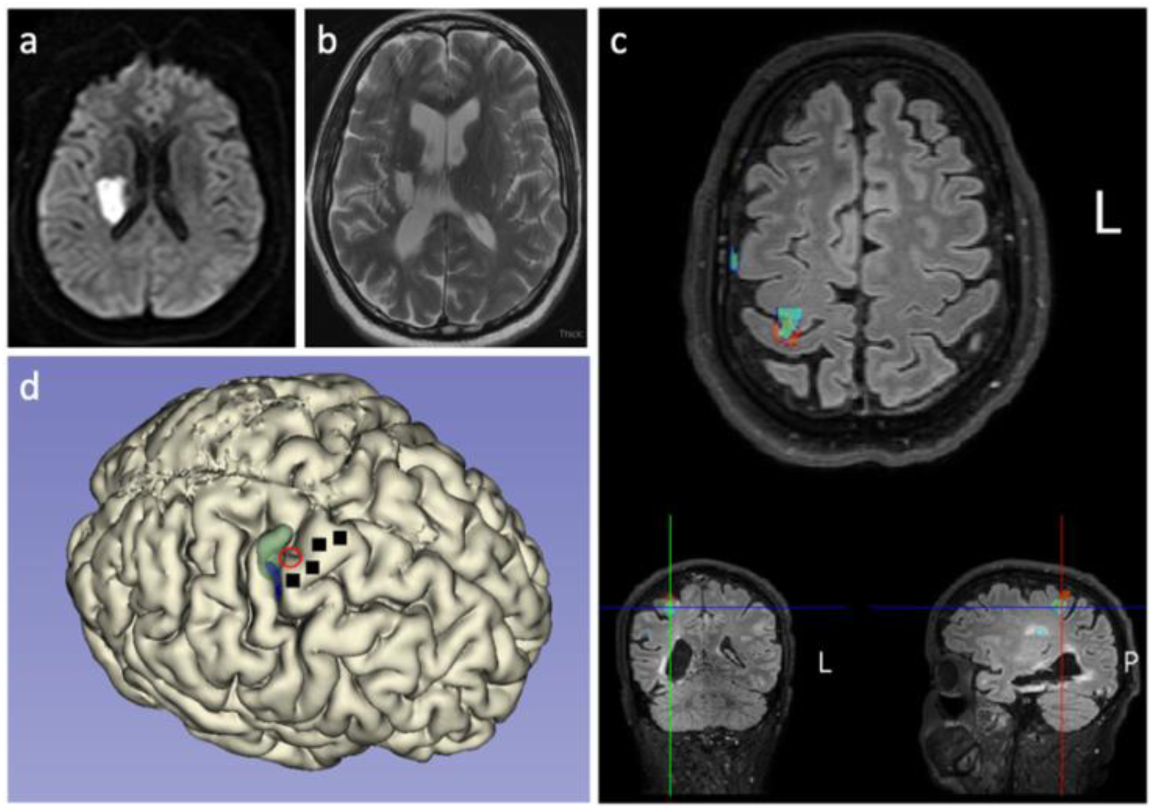
Neuroimaging results. Diffusion sequence when the acute stroke occurred; diffusion restriction is evident in the right lentiform nucleus and adjacent white matter (a). T2-weighted MRI two years later shows areas of encephalomalacia and relative ventriculomegaly (b). Functional neuroimaging revealed a hot spot of activation, indicated by a red circle, in the depth of the central sulcus along the ‘hand knob’ area of the precentral gyrus (c). A three-dimensional reconstruction of the participant’s cortical surface derived from MRI with imagined left hand movement centroid of activity indicated by the red circle (d). Green shading indicates an area responsive to sensory stimulation of the left hand. Black squares indicate microelectrode arrays.

### Neural recordings

Well-delineated single units were recorded from 87 of the 256 channels (Fig. 4). Neural activity correlated with actual and attempted movements in both the paretic left arm in addition to the intact right arm. The discharge rate of various units appeared to correlate with specific residual actions, including the wrist extension that gradually developed in the course of the three-month duration (Fig. 5). By taking the spike counts recorded at each channel every 200 milliseconds and running them through a leaky integrator^21^, and then summing these leaky integrator outputs across all channels, we were able to visualize the cumulative cross-array firing rate activity in comparison to forearm electromyographic activity (Fig. 6). Of the 256 electrodes, in each session, we identified 40 channels that were eventually used for neural decoding. These channels were used for extraction of neural features that coded for hand and elbow flexion and extension. Two main hand open-close decoding approaches were used: 1) A discrete two-state classifier based on a 1-dimensional linear filter continuous output; 2) a “training-less” threshold crossing approach with a rolling baseline normalization.

**Figure 4.**
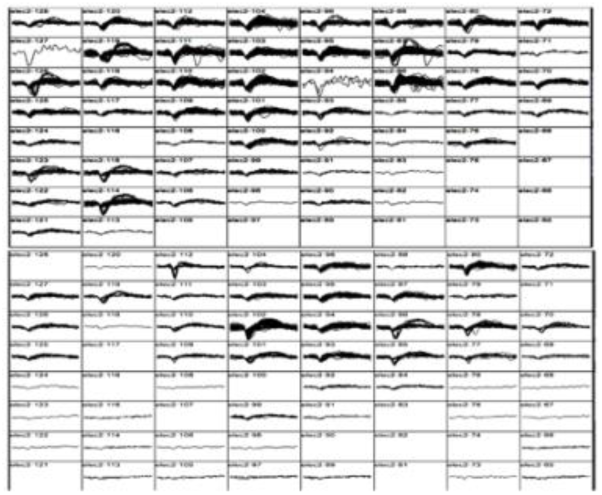
Action potential waveforms. Snapshot of action potential waveforms recorded from two of the arrays.

**Figure 5.**
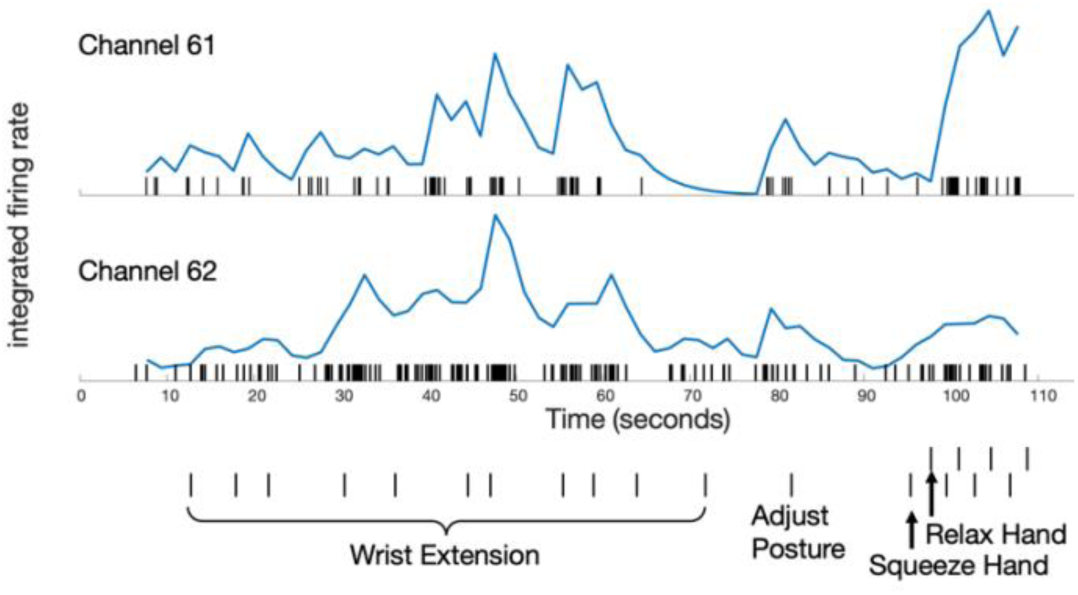
Neuronal activity correlated with performed movements in the paretic limb. Over a 110-s period, the participant was asked to perform a series of left limb movements (described on abscissa). Verbal movement instructions indicated by hash marks. Rasters indicate the time of each action potential. Normalized, integrated firing rates appear beneath each raster, derived by a ‘leaky integrator’ equation in ^21^; normalization achieved by dividing by the maximum integrated firing rate from each unit’s spike train over the time period displayed. The top unit (channel 61) is more active for hand squeezing than wrist extension, relative to the bottom, simultaneously recorded unit (channel 62). The participant performed all movements: such motions required effort and he was unable to engage a consistent level of activity for each cue and exhibited a variable reaction time. The participant was easily fatigued, requiring him to take a break and adjust posture.

**Figure 6.**
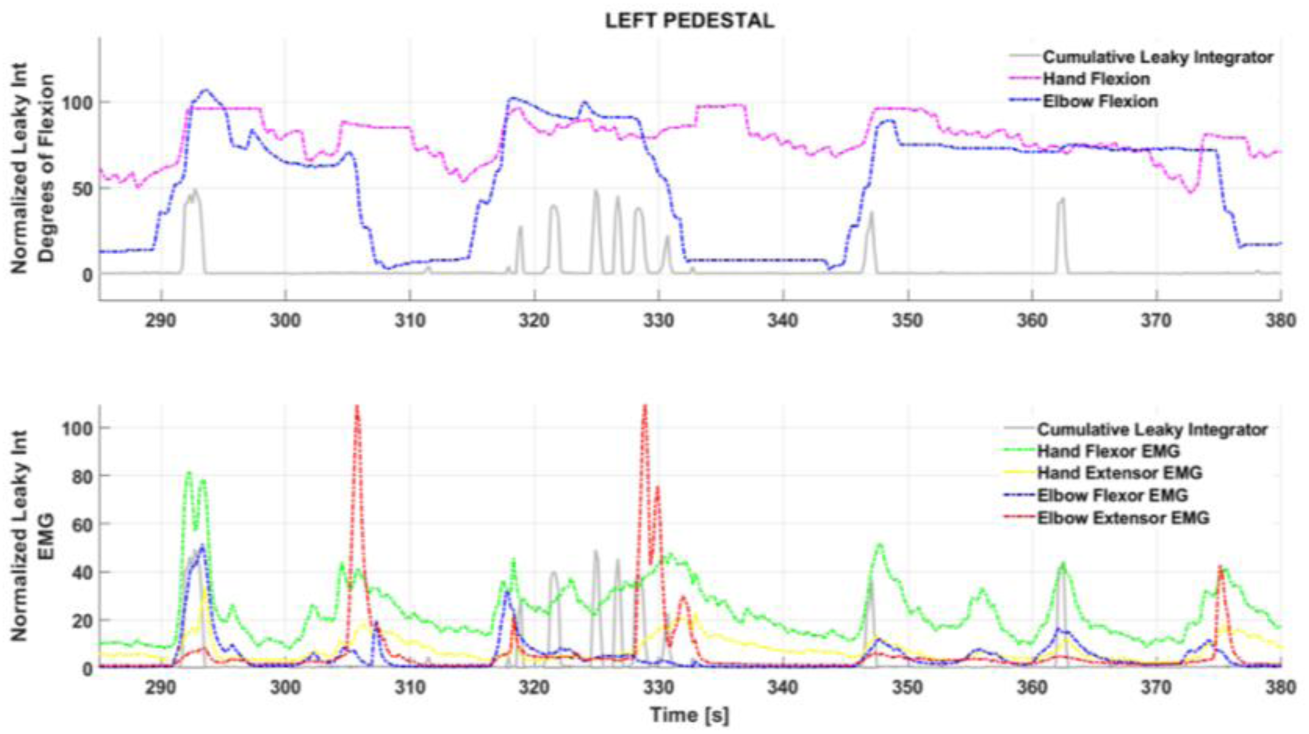
Cumulative, integrated spike activity across channels fluctuating with joint position (top) and residual left forearm electromyographic activity (bottom). The summed spike activity across channels and run through a leaky integrator ^21^ appeared to fluctuate with specific residual actions in the left upper extremity. Proximal residual activity generated a normal-appearing pattern: as seen between 290 and 310 seconds in the bottom panel, biceps (blue trace) and triceps (red trace) activity alternate. In the distal upper extremity, however, wrist flexor (green) and wrist extensor (yellow) trace, tend to occur together in an abnormal synergy; also, wrist flexor activity is abnormally synergistic with biceps activity (an abnormal flexor synergy). The summed, integrated spiking activity across channels appears to covary with wrist flexor activity.

### Orthosis control

The left upper extremity score on the Action Research Arm Test (ARAT)^19^ was 0 without the orthosis on, 5 using the orthosis under myoelectric control, and 10 using the orthosis under direct brain-control. In one component of the Jebsen-Taylor standardized test of hand function^22^, the goal is to pick up and move 5 cans, one at a time, a few inches away forward on the table (normal times are 3.23 seconds for empty soup cans in subtest 6, and 3.30 seconds for full cans in subtest 7). Because the design of the hand orthosis precluded the ability to grasp a soup can (i.e., the brace only supports the thumb and next two digits), the participant performed variations on the test. It took the participant 146 seconds to pick up, move and release 5 pill bottles using myoelectric control, and 95 seconds to perform the identical task under BCI control. Another task was to hold an object in the right hand (e.g., a stress ball or a whiteboard eraser) and place it into the paretic left hand, and then extend the left arm down towards the floor and drop the object into a bin; this process was then repeated 5 times in a row. Both these tasks were performed with the participant seated. On two trials of this pick-up-and-drop-5 in myoelectric mode, the participant’s completion times were 128 and 222 sec/trial; seconds; on five trials of the same task in BCI mode, times were 81, 106, 137, 214 seconds. In addition to measuring the total time to perform these grasp-move-release tasks, we also quantified the time it took to release an object once the hand was in the target position. Hand release times were faster under BCI control than myoelectric control (*p=0*.*04*, two-sample t-test).

### Motor outcomes

Motor measures, performed when the participant was not connected to the BCI or wearing the MyoPro orthosis, were tracked over time and demonstrated that the implantation procedure did not decrease residual strength in the paretic left arm and left leg. In fact, muscle strength increased in the left arm. Whereas serial neurological exams since the time of the stroke demonstrated an absence of voluntary wrist extension or finger extension (0/5 on manual muscle testing, starting two months into the trial, the participant began to consistently exhibit voluntary wrist extension against gravity (3/5), and on a few occasions was able to voluntarily extend the fingers slightly (2/5)^23^. One month prior to the device implantation, the Fugl-Meyer upper extremity score was 30 (out of a maximum of 66) for the left upper extremity; this increased to a score of 36 four weeks after the two Multiports were implanted, and a score of 38 seven weeks post-implantation^24^. Although the participant did not receive botulinum toxin injections, or receive any type of anti-spasticity medication, during the clinical trial, spasticity gradually decreased with time as reflected in gradually decreasing numbers on serial measurements of the modified Ashworth scale for spasticity for passive flexion and extension movements of the fingers, wrist, and elbow, along with internal and external rotation of the shoulder^25^.

## Discussion

This pilot trial demonstrated that ensemble single unit activity remains active in ipsilesional cerebral cortex overlying chronic subcortical stroke. To our knowledge, this is the first report of intracortical recordings in ipsilesional cerebral cortex for a stroke above the mesencephalon. The trial established that single neuron, movement related activity can be decoded used to control a powered orthosis that restored functionally useful voluntary upper extremity movement. Importantly, this brain-computer interface system can be used simultaneously with residual intact movement, in particular in a limb with a gradient of intact to absent voluntary movement, as is common following cerebral strokes. While myoelectric approaches based upon wrist flexion did enable voluntary hand opening, this approach triggered increased muscle tone that subsequently slowed orthosis use (as the motors were opposing the abnormal tone): the BCI control mode essentially bypassed this issue and allowed motors to operate more smoothly and quickly. Electromyographic recordings demonstrated that while the participant did continue to engage wrist flexors during BCI control, the amplitude was decreased from abnormally elevated levels to more normal amplitudes.

This trial was not intended to restore voluntary motor control in the hemiparetic upper extremity in the absence of any device use, but even so, we found that strength improved, and spasticity decreased. This suggests that the implantation of four arrays into ipsilesional cortex did not exacerbate pre-existing hemiparesis (i.e., it did not worsen hand or arm weakness); indeed, after the intervention hand functions improved. One potential explanation for the unexpected improvements in voluntary wrist and finger extension is mass practice. Another, more speculative, explanation for the participant’s improved forearm function is that the daily exercise of ipsilesional cortical activity for BCI-orthosis control, promoted a plasticity driven response to either normalize or compensate for abnormal motor synergies.

Although the limited number of trials on various tasks reduced statistical power to compare myoelectric to BCI control, qualitatively there appeared to be a trend of faster control in the BCI mode. This may be due to the fact that triggering orthosis action from direct cortical recordings does not activate abnormal forearm synergies in the same manner that myoelectric control appears to. Spasticity may represent abnormal plasticity and loss of corticoreticular facilitation of the medullary inhibition center leading to decreased inhibition from the dorsal reticulospinal tract on the spinal stretch reflex: the medial reticulospinal and vestibulospinal tracts are unopposed leading to stretch reflex hyperexcitability^26^. In the myoelectric mode, where hand closing is triggered by activation of residual wrist flexors, this hyperexcitability is inevitably triggered such that the orthosis motors have to ‘fight harder’ to open the hand, slowing that process. In the BCI mode, even if residual wrist flexor and extensor activity are engaged, it is to lesser degree such that abnormal tone is not elevated, and the orthosis motors can more easily and rapidly achieve hand actions.

This pilot study implies that usable control signals are present in ipsilesional cerebral cortical activity. To be clinically scalable, future devices must be fully implantable to minimize infection risk and allow mobility. With the advent of fully implantable BCI (i.e., no percutaneous connectors, ^27,28 29^), a wider range of stroke survivors could benefit. An option that may gain even wider clinical adoption would be to couple direct cortical control to implantable functional electrical stimulation in the paretic arm, as has been demonstrated in at least one person with chronic stroke^10^. Direct cortically driven peripheral muscular stimulation may have both rehabilitative^30^ and direct functional benefits if deployed continuously in daily life. Fully implantable brain-computer interfaces may represent a medical device opportunity to help stroke patients break through their plateau in recovery and to achieve greater functional independence.

## Data Availability

Data sharing agreements are available and can be requested by contacting the first author.

## Ethics Committee Approval

This research was approved, and remains in approved status, by the Institutional Review Board of Thomas Jefferson University, protocol number 17D.459.

## Acknowledgements

The authors would like to thank the following people for their assistance and input: Erica Jones, Shivayogi Hiremath, Christopher Thompson, Carlos Vargas-Irwin, John Donoghue, Nicholas Hatsopoulos, David Weisman, Kristofer Feeko, M.J. Mulcahey, Joseph Tracy, Diana Tzeng, Daniel Graves, Ashly Parekh, Joely Mass, Thomas J. Kelly, IV, Stephen Valverde, Allison Weiss, Shaista Alam, Robin Dharia, Elan Miller, Lisa Bowman, Rodney Bell, Michael Sperling, and the participant and his mother.

## References

1. Katan, M. & Luft, A. Global Burden of Stroke. Semin. Neurol. (2018). doi:10.1055/s-0038-1649503

2. Vos, T. et al. Global, regional, and national incidence, prevalence, and years lived with disability for 310 diseases and injuries, 1990–2015:a systematic analysis for the Global Burden of Disease Study 2015. Lancet (2016). doi:10.1016/S0140-6736(16)31678-6

3. Centers for Disease Control. Prevalence of stroke--United States, 2006-2010. MMWR. Morbidity and mortality weekly report 61, (2012).

4. Kwakkel, G., Kollen, B. J., Van der Grond, J. V. & Prevo, A. J. H. Probability of regaining dexterity in the flaccid upper limb: Impact of severity of paresis and time since onset in acute stroke. Stroke (2003). doi:10.1161/01.STR.0000087172.16305.CD

5. Milekovic T, Sarma AA, Bacher D, Simeral JD, Saab J, Pandarinath CS. B., Blabe C, Oakley EM, Tringale KR, Eskandar E, Cash SS, Henderson JM, S. K. & Donoghue JP, H. L. Stable long-term BCI-enabled communication in ALS and locked-in syndrome using LFP signals. J Neurophysiol (2018). doi:10.1152/jn.00493.2017

6. Klaes, C. et al. A cognitive neuroprosthetic that uses cortical stimulation for somatosensory feedback. J. Neural Eng. 11, 056024 (2014).

7. Collinger, J. L. et al. High-performance neuroprosthetic control by an individual with tetraplegia. Lancet 381, 557–564 (2013).

8. Bhagat, N. A. et al. Design and optimization of an EEG-based brain machine interface (BMI) to an upper-limb exoskeleton for stroke survivors. Front. Neurosci. 10, (2016).

9. Page, S. J., Hill, V. & White, S. Portable upper extremity robotics is as efficacious as upper extremity rehabilitative therapy: a randomized controlled pilot trial. Clin. Rehabil. 27, 494–503 (2013).

10. Knutson, J. S. et al. Implanted neuroprosthesis for assisting arm and hand function after stroke: A case study. J. Rehabil. Res. Dev. 49, 1505 (2012).

11. Knutson, J. S. et al. Adding contralaterally controlled electrical stimulation of the triceps to contralaterally controlled functional electrical stimulation of the finger extensors reduces upper limb impairment and improves reachable workspace but not dexterity: A randomized. Am. J. Phys. Med. Rehabil. (2020). doi:10.1097/PHM.0000000000001363

12. Wisneski, K. J. et al. Unique cortical physiology associated with ipsilateral hand movements and neuroprosthetic implications. Stroke 39, 3351–3359 (2008).

13. Knutson, J. S. et al. Implanted neuroprosthesis for assisting arm and hand function after stroke: a case study. J. Rehabil. Res. Dev. 49, 1505–16 (2012).

14. Bundy, D. T. et al. Using ipsilateral motor signals in the unaffected cerebral hemisphere as a signal platform for brain-computer interfaces in hemiplegic stroke survivors. J. Neural Eng. 9, 036011 (2012).

15. Vargas-Irwin, C. E. et al. Watch, imagine, attempt: Motor cortex single-unit activity reveals context-dependent movement encoding in humans with tetraplegia. Front. Hum. Neurosci. (2018). doi:10.3389/fnhum.2018.00450

16. Hochberg, L. R. et al. Reach and grasp by people with tetraplegia using a neurally controlled robotic arm. Nature 485, 372–375 (2012).

17. Suner, S., Fellows, M. R., Vargas-Irwin, C., Nakata, G. K. & Donoghue, J. P. Reliability of signals from a chronically implanted, silicon-based electrode array in non-human primate primary motor cortex. IEEE Trans. Neural Syst. Rehabil. Eng. 13, 524–541 (2005).

18. Rousche, P. J. & Normann, R. A. A method for pneumatically inserting an array of penetrating electrodes into cortical tissue. Ann. Biomed. Eng. (1992). doi:10.1007/BF02368133

19. Yozbatiran, N., Der-Yeghiaian, L. & Cramer, S. C. A standardized approach to performing the action research arm test. Neurorehabil. Neural Repair (2008). doi:10.1177/1545968307305353

20. Taylor, N., Sand, P. L. & Jebsen, R. H. Evaluation of hand function in children. Arch. Phys. Med. Rehabil. (1973).

21. Fetz, E. E. & Baker, M. A. Operantly conditioned patterns on precentral unit activity and correlated responses in adjacent cells and contralateral muscles. J. Neurophysiol. (1973). doi:10.1152/jn.1973.36.2.179

22. Jebsen, R. H., Taylor, N., Trieschmann, R. B., Trotter, M. J. & Howard, L. A. An objective and standardized test of hand function. Arch. Phys. Med. Rehabil. (1969).

23. Martin, E. G. & Lovett, R. W. A Method of Testing Muscular Strength in Infantile Paralysis. J. Am. Med. Assoc. (1915). doi:10.1001/jama.1915.02580180016006

24. Fugl-Meyer, A. R., Jääskö, L., Leyman, I., Olsson, S. & Steglind, S. The post-stroke hemiplegic patient. 1. a method for evaluation of physical performance. Scand. J. Rehabil. Med. (1975). doi:10.1038/35081184

25. Ashworth, B. PRELIMINARY TRIAL OF CARISOPRODOL IN MULTIPLE SCLEROSIS. Practitioner (1964).

26. Li, S. Spasticity, motor recovery, and neural plasticity after stroke. Frontiers in Neurology (2017). doi:10.3389/fneur.2017.00120

27. Yin, M. et al. Wireless neurosensor for full-spectrum electrophysiology recordings during free behavior. Neuron (2014). doi:10.1016/j.neuron.2014.11.010

28. Musk, E. An Integrated Brain-Machine Interface Platform With Thousands of Channels. J. Med. Internet Res. (2019). doi:10.2196/16194

29. Kohler, F. et al. Closed-loop interaction with the cerebral cortex: a review of wireless implant technology§. Brain-Computer Interfaces (2017). doi:10.1080/2326263X.2017.1338011

30. Olsen, S. et al. Peripheral Electrical Stimulation Paired With Movement-Related Cortical Potentials Improves Isometric Muscle Strength and Voluntary Activation Following Stroke. Front. Hum. Neurosci. (2020). doi:10.3389/fnhum.2020.00156

